# Continuing to be Cautious: Japanese Contact Patterns during the COVID-19 Pandemic and their Association with Public Health Recommendations

**DOI:** 10.1101/2025.04.17.25326019

**Authors:** Tomoka Nakamura, Ryo Kinoshita, Akira Endo, Katherine E. Atkins, Hitoshi Oshitani, Yoko Ibuka, Motoi Suzuki, Koya Ariyoshi, Kathleen M. O’Reilly

**Author notes:** Corresponding author: Tomoka Nakamura.

## Abstract

Despite implementing no lockdowns and having a large elderly population, Japan had a low mortality rate due to COVID-19 compared to Europe and North America. The extent to which policies impacted person-to-person contact remains unclear. In this study, we examined changes in contact patterns and their association with behaviors and governmental recommendations in Japan during the pandemic.

Ten social contact surveys were conducted between 2021 and 2023 reaching over 1500 participants per survey in Osaka and Fukuoka prefectures where governmental recommendations were first implemented due to high COVID-19 incidence. Their contact patterns were assessed through their demographic characteristics, COVID-19 vaccination status, and individual disease mitigation measures. Generalized linear models were used to identify factors associated with increased contacts.

The mean number of contacts during the pandemic declined by at least 49.8% (8.2 weekday contacts and 6.0 weekend contacts per individual, adjusted by age and sex) compared to a study conducted prior to 2020. Weekdays, occupation, larger household sizes, and mask wearing were associated with a higher number of contacts. The frequency and duration of contacts were negatively associated with the issuance of COVID-19 governmental measures, yet the relative change in contacts was not as prominent as pre- and post-lockdown situations in the United Kingdom.

There was a gradual increase in contacts with time and less strict public health recommendations. Yet, contacts that did not increase with uptake of COVID-19 vaccination and continuous mask wearing depict cautious behavior across the survey population during the pandemic and into 2023. These results are in contrast with European countries where contacts largely increased among vaccinated individuals compared to the non-vaccinated. Social contacts are country and context specific, highlighting the need for data collection across different communities.

## Background

The reported cases and deaths due to coronavirus disease 2019 (COVID-19) in Japan have been much lower compared to Western countries such as Europe and North America. For comparison, as of 28 June 2023, there were 74,694 cumulative deaths in Japan in a total population of 125.5 million (59.5 deaths per 100,000) while there was a total of 227,524 deaths in the United Kingdom with a population approximately half of Japan (337.9 deaths per 100,000) (1). Japan is also unique having the second largest aging population in the world, after Monaco, (29.9% of the total population who are 65 years and above) (2) who are at a higher risk of COVID-19 mortality. Throughout the pandemic from 2020 to early 2022, cumulative confirmed COVID-19 cases (adjusted by population size) were reported 10 to 15 times higher in the United States and United Kingdom compared to Japan (1). Several factors, such as timing of government regulations and behavioral change interventions, have been discussed as potential determinants of a successful epidemic control (3).

Apart from strict international border control policies, Japan relatively has had less strict rules compared to many other countries. Though public health emergency declarations (EDs) with different levels of strictness were issued, neither lockdowns nor curfews were implemented. National school closure due to COVID-19 regulation spanned for at least three weeks in March 2020, but most schools reopened between April and May 2020 (4). Yet, there were key messages that continued to be addressed to the public, such as avoiding the “3Cs” which stands for settings that are closed, crowded and close contact (5). Reducing person-to-person contact has been one of the key tactics in epidemic control as it directly shapes the risk of transmission of respiratory viruses (6).

To quantify these contact patterns, mobile phone data (7) and synthetic contact matrices (8) have been used. During the COVID-19 pandemic in Japan, mobile device data was utilized to investigate how governmental interventions could have impacted mobility (9,10). Social contact survey data is another important input for epidemiological and mathematical models of infectious diseases. It has its advantages over mobile device data as it can capture changes in contact patterns with respect to age and sex of both survey participants and their contacts. Additionally, disease mitigation measures, such as vaccination and handwashing, can be linked with contact data. Social contact surveys have been well-utilized prior to the pandemic, such as POLYMOD in Europe (11) and in Japan (12,13). They showed that contact patterns are highly dependent on age, gender, household size and day of the week (14). These studies provide a baseline of contact patterns prior to any physical distancing or public health related measures.

To elucidate the changes in contact patterns relevant in the transmission of SARS-CoV-2 in Japan, we conducted 10 repeated cross-sectional surveys from 2021 to 2023 (Figure 1a, 1b). In this paper, we aim to describe the changes in social contacts and other behaviors, such as hand hygiene and mask wearing, during the pandemic and into 2023. As different levels of EDs were issued and lifted with time, we evaluated the association of these governmental measures on social contacts. We also present a statistical model that explores individual characteristics and behavior that impact the frequency of contacts.

**Fig 1a:**
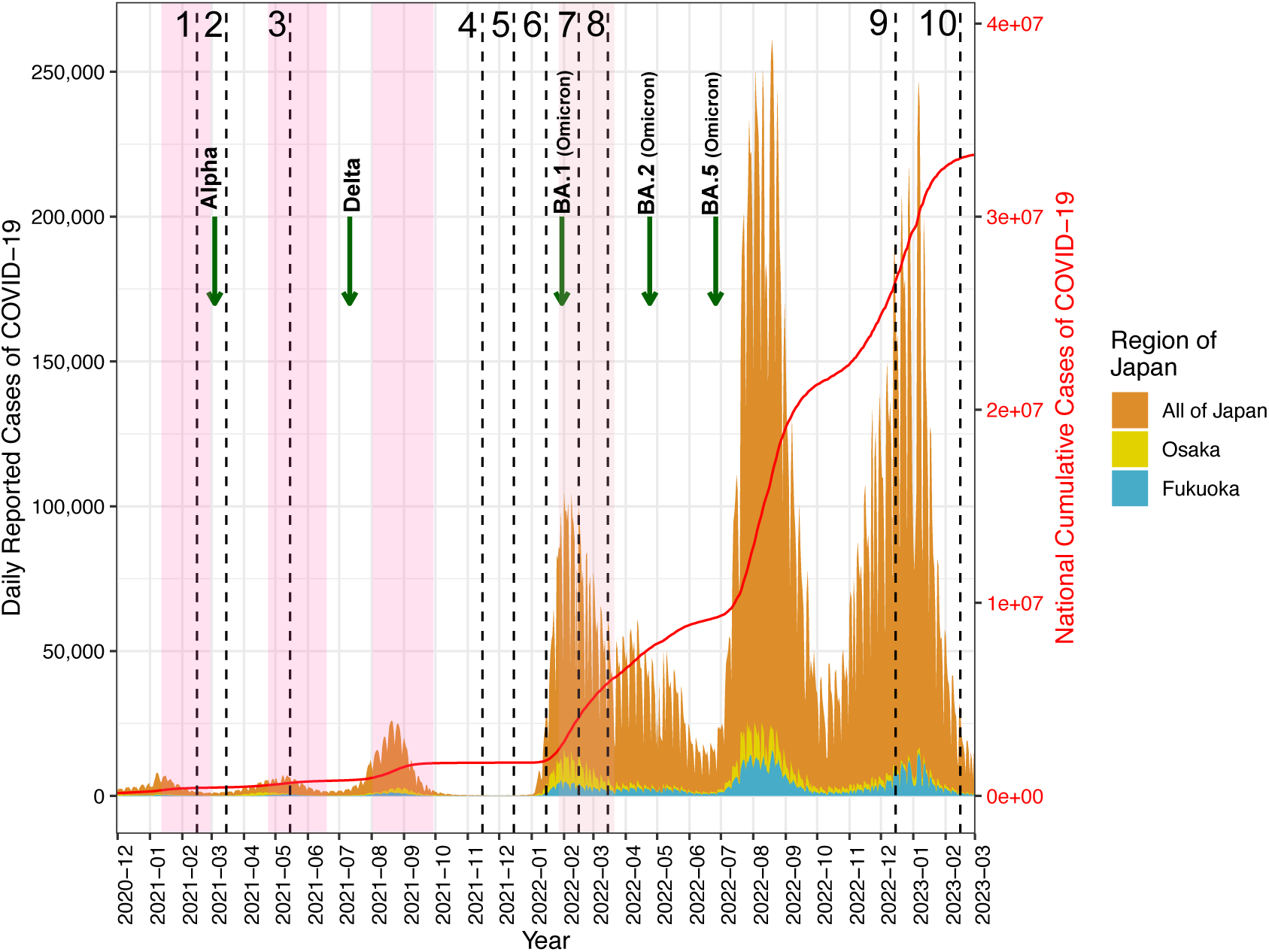
Number of daily and cumulative reported COVID-19 cases in Japan with the timing of the social contact surveys in this study and introduction of SARS-CoV-2 variants. The dashed vertical lines indicate each time point when the contact surveys were conducted beginning from February 2021 to February 2023. The darker pink rectangles indicate the periods when emergency declarations were issued in Osaka and Fukuoka. The lighter pink rectangle indicates the period of semi-emergency declaration that was issued in Osaka and Fukuoka. In **Fig 1a**, the green arrows indicate the beginning of the transmission period when at least 50% of the genome sequenced COVID-19 cases was due to specific variants of SARS-CoV-2. In **Fig 1b**, the purple arrow indicates the start of the Tokyo Summer Olympics in 2021. Navy arrows indicate national holidays in Japan. Light green arrows indicate the dates of mass vaccination that occurred sequentially based on priority groups.

**Fig 1b:**
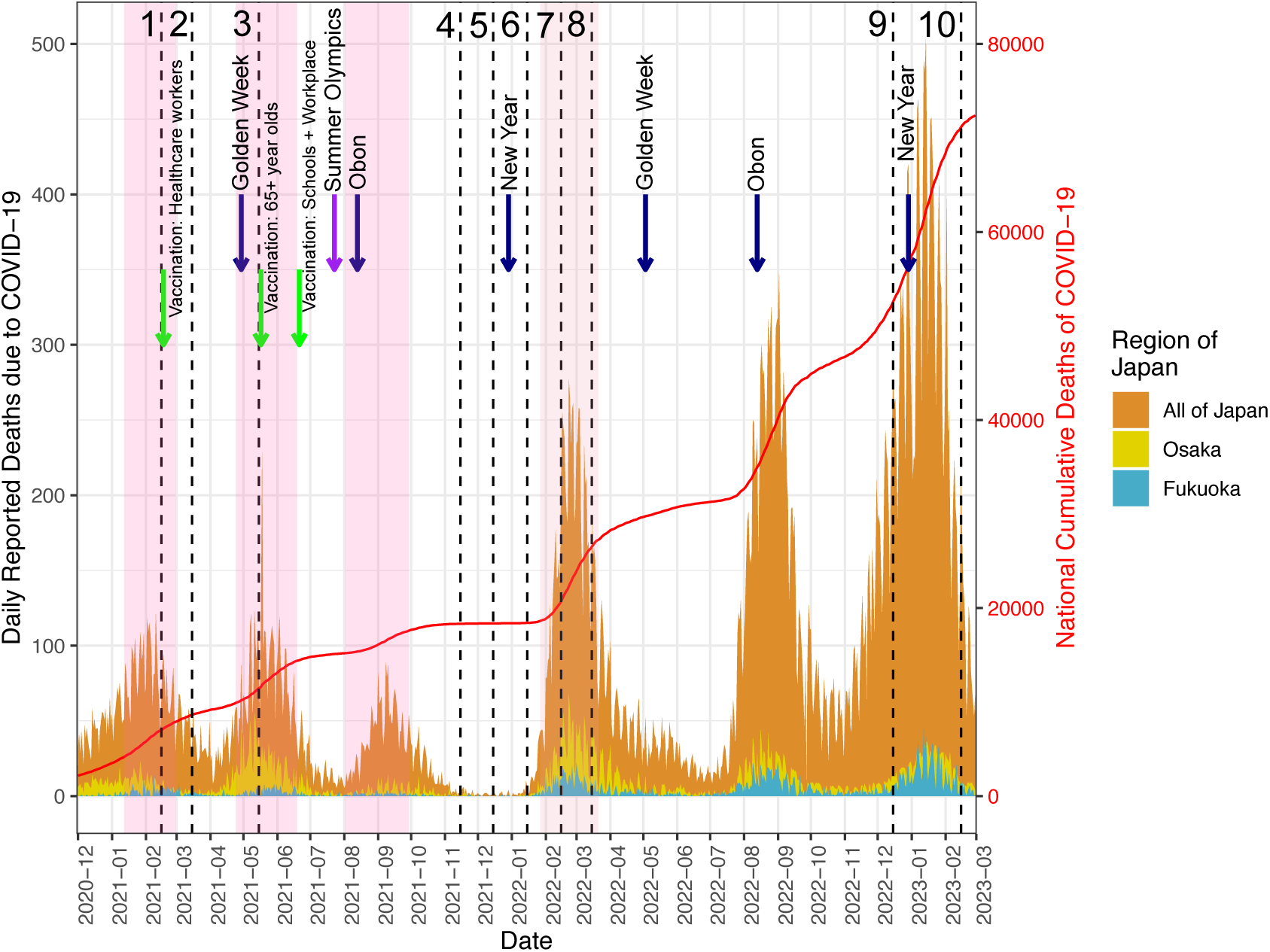
Number of daily and cumulative reported COVID-19 deaths in Japan with the timing of the social contact surveys in this study and key events related to COVID-19.

## Methods

### Survey design

Ten contact surveys, in which the participants were asked to report their individual characteristics and contact patterns, were conducted in Osaka and Fukuoka prefectures, Japan in 2021–2023 (Figure 1). Osaka prefecture is located on the west of mainland Japan and has the third highest population next to Tokyo and Kanagawa prefectures (15). Fukuoka prefecture is on Kyushu island, which is south of mainland Japan, and has the ninth highest population (15). In 2021, three EDs were issued across Japan including Osaka and Fukuoka during which two surveys (#1 and 3, Figure 1) were conducted. During the height of the BA.1 (omicron) transmission in 2022, a semi-ED was issued, which consisted of a less strict recommendation than an ED (Suppl. Table 1) (16). Two surveys (#7 and 8, Figure 1) were conducted during this time. Six surveys (#2, 4, 5, 6, 9, and 10, Figure 1) were conducted when ED was absent with the last survey in February 2023. The timing of the surveys was semi-strategic; they were planned during expected changes in contact patterns and funding availability.

Survey participants were recruited by a Japanese online survey company (F-press). Participants included anyone at least 18 years old residing in either Osaka or Fukuoka prefecture who agreed to participate through written informed consent. Children under 18 years old participated with the consent of their guardians/parents who recorded their information on their behalf. Every individual in each household can fill out the survey as a participant, and all these individuals can be linked by household. The number of participants per survey was a minimum of 1,500, powered to detect a difference in contact numbers of 2.5 between pairs of observations with a 90% power and 5% Type I error and 20% loss to follow-up. The survey participants were compensated 300 Japanese yen per survey and were able to participate in as many surveys as they opted for between 10 February 2021 and 12 February 2023.

The survey was adapted from the UK CoMix study (17–19) in Japanese to capture the daily frequency and type of contacts. In each survey, they recorded their contacts during one weekday and one of the days during the weekend. A “contact” was defined as physical or non-physical: physical contact included handshakes, hugging, kissing, and playing a contact sport while non-physical contact was defined as facing another individual (with or without mask wearing) and exchanging at least three Japanese sentences to each other in a conversation. The participants were asked to record their contacts in a diary format with instructions a week prior to the day when the survey was implemented. They reported their contacts including their age, sex, type of contact (non-physical and/or physical), location of contact, approximate time of contact, and whether the contact was made indoors and/or outdoors (Supp. Table 1). These specified contacts were for the first 10 contacts. Those who reported more than 10 contacts were asked to approximate the number of contacts, location of contacts, and their age categories.

Participants were asked to report their individual characteristics as well as their individual preventive behavior such as frequency of mask wearing, handwashing, and teleworking (Supp. Table 1). They were asked COVID-19 related questions including their history of having tested PCR positive for COVID-19, their concern towards getting infected with COVID-19 (rank from one to five), and their vaccination status.

### Statistical analysis

For each survey, we calculated the mean and median of contacts during the weekday and weekend, mean and median age, COVID-19 vaccine coverage, frequency of mask wearing, and the proportion of survey participants who tested COVID-19 positive. The population estimates of the mean contacts and vaccination coverage were adjusted by age and sex based on the 2021 October census (15).

We compared our data to contact patterns analyzed by Ibuka et al. (12) as baseline data prior to the COVID-19 pandemic. Their survey methodology was similar to ours; the data included all reported contacts per individual during the week as well as the duration and location of contacts. Their contact surveys were also conducted with an objective to understand influenza-related behaviors which is relevant in the context of respiratory disease transmission. For our study, contact patterns were stratified by weekday and weekend and compared across eight age groups (0-9, 10-19, 20-29, 30-39, 40-49, 50-59, 60-69, and 70+) and across the study period.

Measures of uncertainty in age-specific contact numbers and duration were obtained using the bootstrap; the mean and 95% confidence intervals (CI) were obtained by sampling with replacement for 1000 times. Consistent with previous contact survey studies, we truncated the total number of contacts to avoid a few observations with hyperinflated contact numbers affecting age-specific summary statistics (11,19). We selected a truncation cutoff of 250 through a visual check by plotting the mean number of contacts with a range of cutoff points using a Weibull distribution. (Supp. Fig 1).

To investigate factors associated with the reported number of contacts (February 2023 survey only), we used a multivariable regression assuming a Weibull distribution with log link function. This distribution was selected to address the coefficient of variation in reported contacts and fit the right skewed distribution of contacts better than a negative binomial distribution. The dependent variable was the reported contacts, and 15 variables were selected from a list of variables (Supp. Table 1) as the model covariates based on a hypothesis-driven approach. The model was referenced on a 40-49 year old individual who lives with two others works as a company employee at their workplace, and fully vaccinated against COVID-19 with at least 3 or more doses. The participants’ age was retained in the model as it could be a potential confounder. All other covariates that were selected in the multivariable model were associated (*p* < 0.05) with the number of contacts in an univariable model. The multivariable model was developed using both forward and backward stepwise selections until the Akaike Information Criterion (AIC) had no further improvement. The incidence rate ratios calculated from the regression model are referred here as contact rate ratios (CRR) where the relative mean number of contacts per day is compared to the reference of each covariate. Model fit was examined with residual plots and comparing predicted vs. observed data. When analyzing summary statistics of contacts across the various time points and age categories, the Kruskal-Wallis test was used. When comparing two nested generalized linear models to test the significance of a variable, likelihood ratio test was used.

All statistical analyses were done in R version 4.0.3, using the packages “MASS”, “survival”, “tidyverse”, “dplyr”, “reshape2”, “scales”, “rstatix”, “ggplot2”, and “cowplot.”

## Results

### Comparison of contact survey participants with national data

A range of 1513 to 1721 participants recorded their contact patterns between February 2021 and February 2023 in Fukuoka and Osaka prefectures, Japan (Table 1). Depending on the survey, the mean age of the participants was between 45 and 48 years old. Between 52 to 54% of the participants were female. As of the 2021 national census, the mean age of the Japanese population was 48.4 and 51.3% being female (15). COVID-19 vaccine coverage among the survey participants increased with time. In Japan, healthcare workers were first vaccinated in February 2021, followed by the older population (≥65 years) from March 2021, and the rest of the population from June 2021 (20). Vaccine coverage reported in February 2023 was slightly higher among the survey participants compared to the national vaccine coverage as of February 2023 (21). Compared to national vaccine coverage reported in January 2023, vaccine coverage across all age groups were slightly higher among the survey participants in February 2023 (Supp. Figure 6). The percent of individuals and/or household members having tested COVID-19 positive increased from 0.3% (5/1721) in February 2021 to 8.7% (136/1569) in February 2023. A serosurvey (N = 5627) conducted between 3^rd^ February and 4^th^ March 2023 showed 35.8% of Osaka residents and 31.3% of Fukuoka residents tested positive with infection-induced antibodies (22). Our survey appropriately approximated the national statistics in terms of demographics but with a slightly higher vaccination coverage and lower percentage of people who tested COVID-19 positive.

**Table 1:**
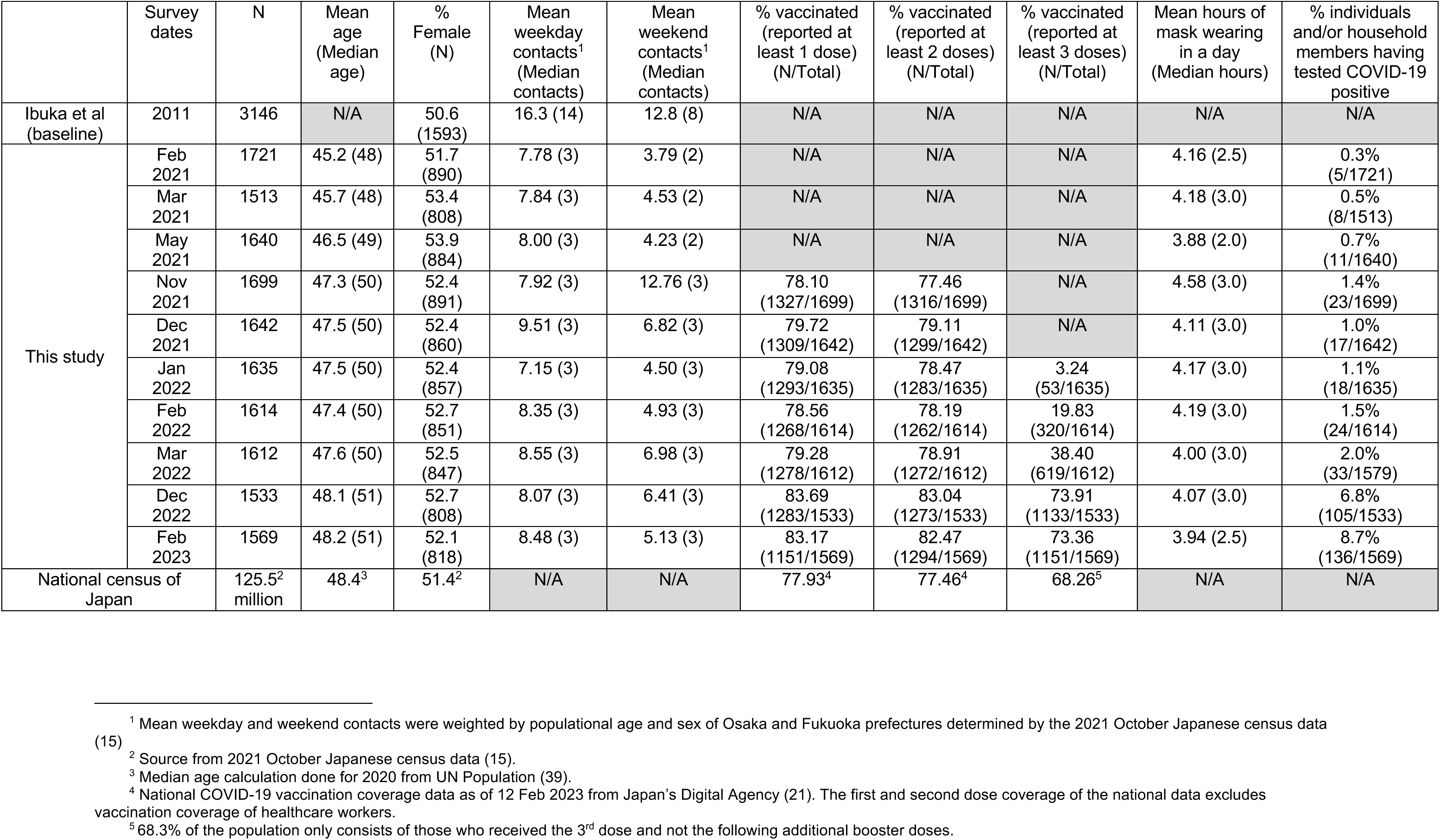
Characteristics of participants across all the contact surveys conducted in Fukuoka and Osaka prefectures from February 2021 to February 2023 compared to pre-pandemic (baseline) contact patterns and the Japanese national census data.

|  | Survey dates | N | Mean age (Median age) | % Female (N) | Mean weekday contacts<sup>1</sup> (Median contacts) | Mean weekend contacts<sup>1</sup> (Median contacts) | % vaccinated (reported at least 1 dose) (N/Total) | % vaccinated (reported at least 2 doses) (N/Total) | % vaccinated (reported at least 3 doses) (N/Total) | Mean hours of mask wearing in a day (Median hours) | % individuals and/or household members having tested COVID-19 positive |
| --- | --- | --- | --- | --- | --- | --- | --- | --- | --- | --- | --- |
| Ibuka et al (baseline) | 2011 | 3146 | N/A | 50.6 (1593) | 16.3 (14) | 12.8 (8) | N/A | N/A | N/A | N/A | N/A |
| This study | Feb 2021 | 1721 | 45.2 (48) | 51.7 (890) | 7.78 (3) | 3.79 (2) | N/A | N/A | N/A | 4.16 (2.5) | 0.3% (5/1721) |
|  | Mar 2021 | 1513 | 45.7 (48) | 53.4 (808) | 7.84 (3) | 4.53 (2) | N/A | N/A | N/A | 4.18 (3.0) | 0.5% (8/1513) |
|  | May 2021 | 1640 | 46.5 (49) | 53.9 (884) | 8.00 (3) | 4.23 (2) | N/A | N/A | N/A | 3.88 (2.0) | 0.7% (11/1640) |
|  | Nov 2021 | 1699 | 47.3 (50) | 52.4 (891) | 7.92 (3) | 12.76 (3) | 78.10 (1327/1699) | 77.46 (1316/1699) | N/A | 4.58 (3.0) | 1.4% (23/1699) |
|  | Dec 2021 | 1642 | 47.5 (50) | 52.4 (860) | 9.51 (3) | 6.82 (3) | 79.72 (1309/1642) | 79.11 (1299/1642) | N/A | 4.11 (3.0) | 1.0% (17/1642) |
|  | Jan 2022 | 1635 | 47.5 (50) | 52.4 (857) | 7.15 (3) | 4.50 (3) | 79.08 (1293/1635) | 78.47 (1283/1635) | 3.24 (53/1635) | 4.17 (3.0) | 1.1% (18/1635) |
|  | Feb 2022 | 1614 | 47.4 (50) | 52.7 (851) | 8.35 (3) | 4.93 (3) | 78.56 (1268/1614) | 78.19 (1262/1614) | 19.83 (320/1614) | 4.19 (3.0) | 1.5% (24/1614) |
|  | Mar 2022 | 1612 | 47.6 (50) | 52.5 (847) | 8.55 (3) | 6.98 (3) | 79.28 (1278/1612) | 78.91 (1272/1612) | 38.40 (619/1612) | 4.00 (3.0) | 2.0% (33/1579) |
|  | Dec 2022 | 1533 | 48.1 (51) | 52.7 (808) | 8.07 (3) | 6.41 (3) | 83.69 (1283/1533) | 83.04 (1273/1533) | 73.91 (1133/1533) | 4.07 (3.0) | 6.8% (105/1533) |
|  | Feb 2023 | 1569 | 48.2 (51) | 52.1 (818) | 8.48 (3) | 5.13 (3) | 83.17 (1151/1569) | 82.47 (1294/1569) | 73.36 (1151/1569) | 3.94 (2.5) | 8.7% (136/1569) |
| National census of Japan |  | 125.5<sup>2</sup> million | 48.4<sup>3</sup> | 51.4<sup>2</sup> | N/A | N/A | 77.93<sup>4</sup> | 77.46<sup>4</sup> | 68.26<sup>5</sup> | N/A | N/A |

<sup>1</sup> Mean weekday and weekend contacts were weighted by populational age and sex of Osaka and Fukuoka prefectures determined by the 2021 October Japanese census data (15)
<sup>2</sup> Source from 2021 October Japanese census data (15).
<sup>3</sup> Median age calculation done for 2020 from UN Population (39).
<sup>4</sup> National COVID-19 vaccination coverage data as of 12 Feb 2023 from Japan's Digital Agency (21). The first and second dose coverage of the national data excludes vaccination coverage of healthcare workers.
<sup>5</sup> 68.3% of the population only consists of those who received the 3<sup>rd</sup> dose and not the following additional booster doses.

### Comparison with pre-pandemic contact patterns

The mean number of contacts during the pandemic reduced by 49.8% during the weekday and by 53.3% during the weekend when compared to pre-pandemic times (12); we report an average of 8.18 weekday contacts and 5.98 weekend contacts per individual. The sample distribution of contacts was right-skewed (Figure 2) where 60.2% of the participants contacted less than five individuals per day during the weekday (and 76.5% during the weekend) in February 2023. The sample distribution of contacts from the earlier surveys showed a similar distribution (Supp. Fig 2). Prior to the pandemic, the distribution of daily reported contacts (N=3146 participants) was also right-skewed and less than 2% of contacts reported zero contacts (12).

**Fig 2:**
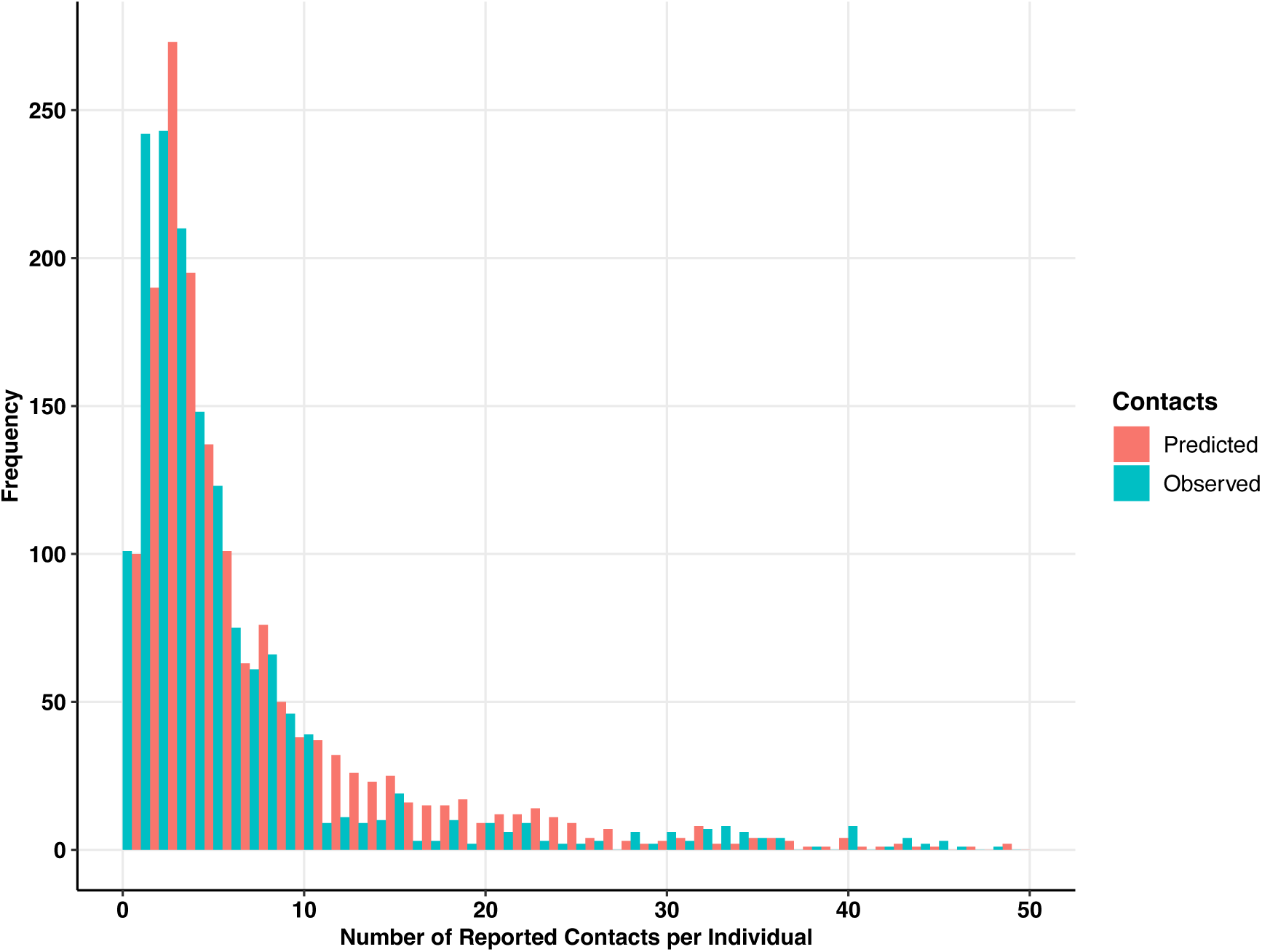
Distribution of contacts reported per individual during the weekday in Fukuoka and Osaka prefectures based on the contact survey conducted in February 2023 among a total of 1569 participants. The blue bars show the observed contacts reported from the contact survey. The pink bars show the predicted contacts based on the multivariable regression model using a Weibull distribution. Sixty participants (3.82%) recorded over 50 contacts.

### Change in contact patterns across a typical week

To test whether there were substantial differences in frequency of contacts across a typical week, two generalized linear models with and without survey time were compared using a likelihood ratio test. Survey time point significantly explained the variability in the mean of contacts during the weekday (chi-squared = 29.33, p-value = 0.00057) (Figure 3a) and during the weekend (chi-squared = 98.79, p-value < 2.2 x 10^-16^) (Figure 3b). Particularly among the 10’s and 40’s, the mean number of contacts increased in calendar time during the weekday and weekend.

**Fig 3:** Frequency (mean) of contacts by age category of participants during the weekday (Fig 3a) and the weekend (Fig 3b) for all surveys conducted (2021-2023) in Fukuoka and Osaka prefectures indicated by each color. The mean and 95% confidence intervals are obtained by bootstrapping.

**Fig 3a:**
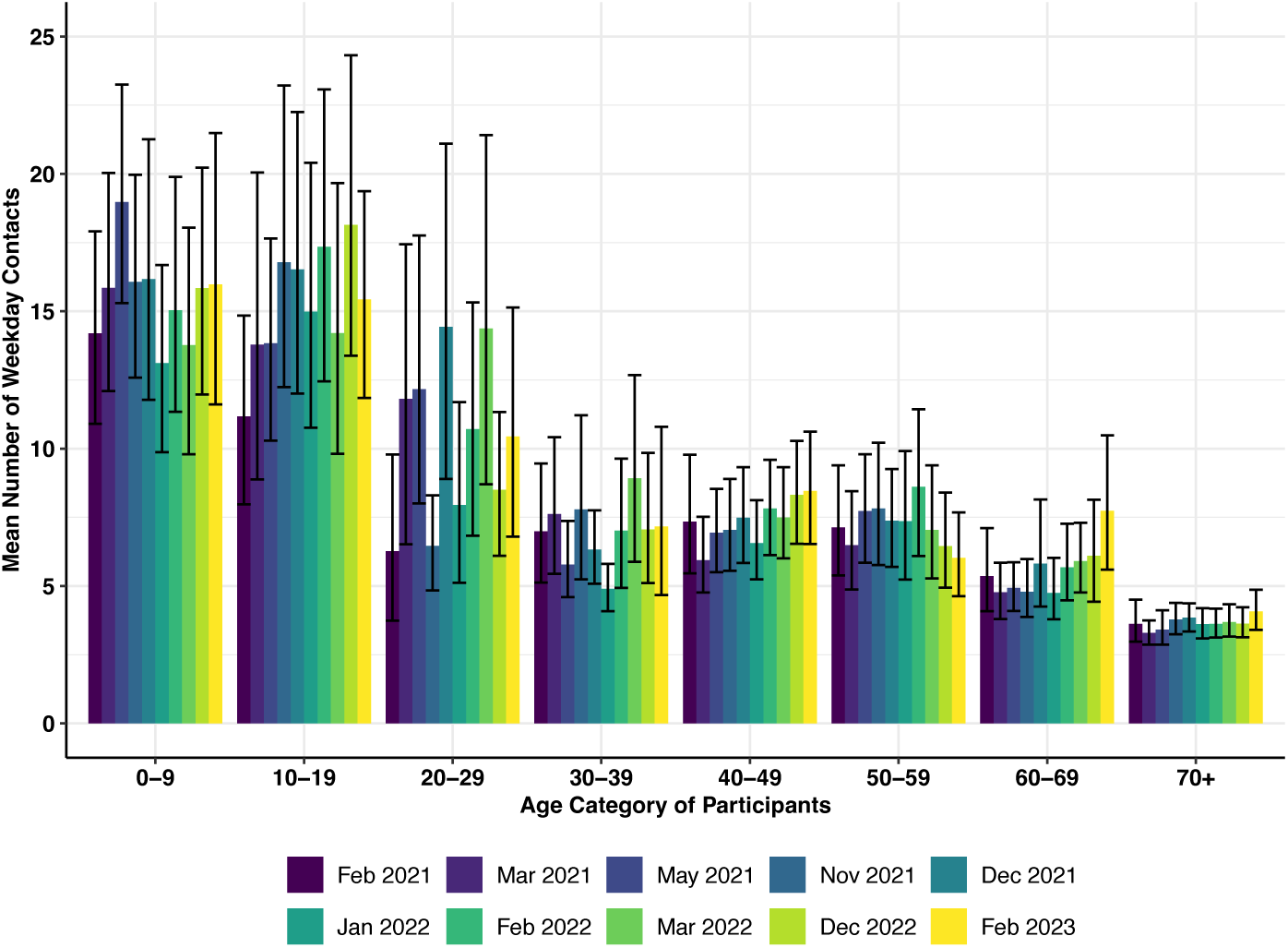
Weekday Contacts 2021-2023

**Fig 3b:**
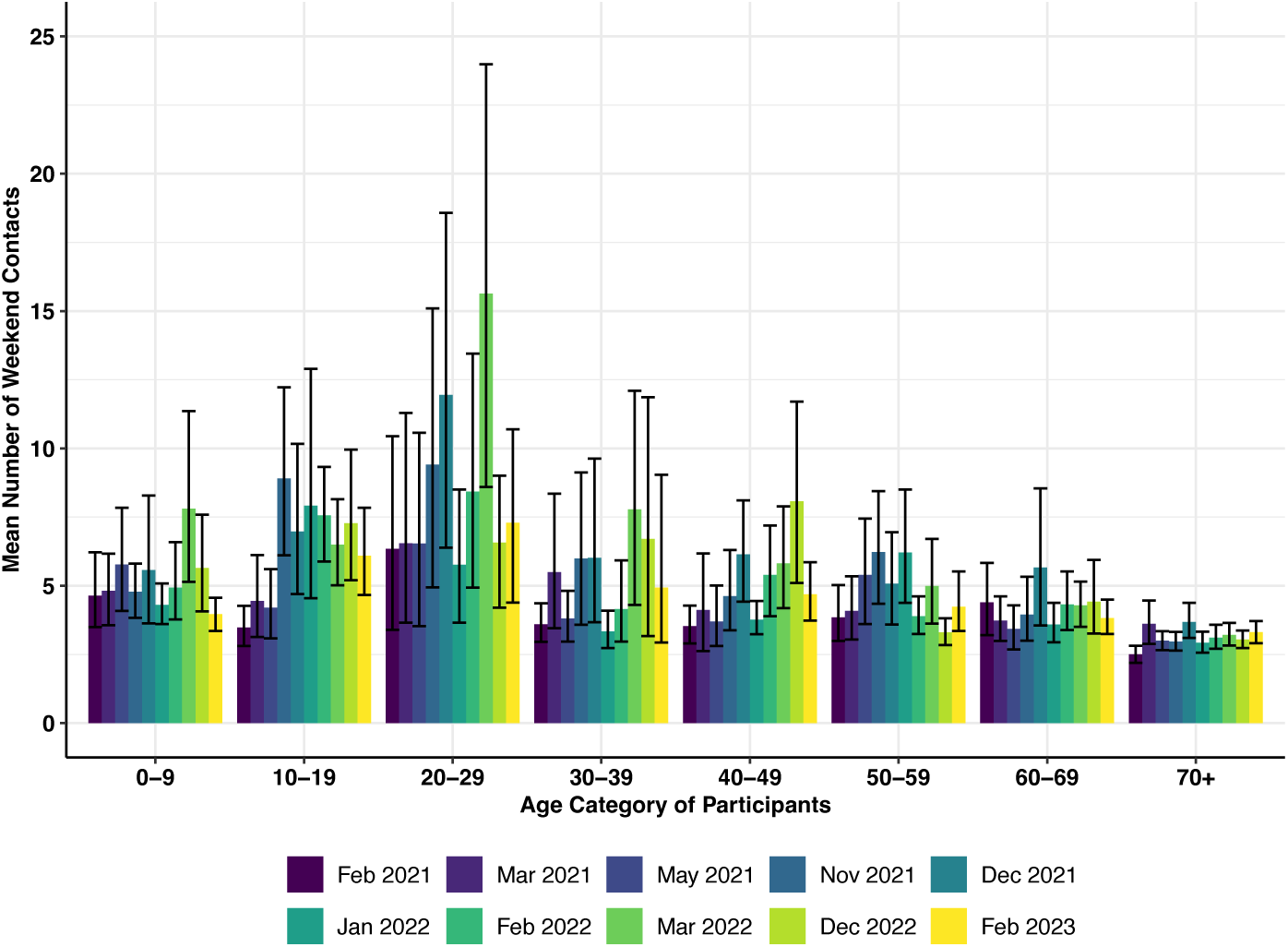
Weekend Contacts 2021-2023

On the contrary, the older population (70+) consistently had low contacts during the weekday and weekend. Particularly during the weekdays, the mean number of contacts across all survey time points (February 2021-February 2023) was significantly lower amongst the 70+ year old compared to the 40’s (chi-squared = 112.65, p-value < 2.2 x 10^-16^). There were clear temporal changes in the duration of contacts in the 40’s during the weekday (chi-squared = 49.47, p-value = 1.35 x 10^-7^) (Figure 4a) and during the weekend (chi-squared = 54.04, p-value = 1.86 x 10^-8^) (Figure 4b). For example, the mean duration of weekday contacts for the 40’s increased from 6.10 hours (95% CI: 5.29-6.85) in February 2021 to 8.63 hours (95% CI: 7.75-9.48) in February 2023. There were similar temporal changes in the duration of contacts amongst the 70+ year old (chi-squared = 21.04, p-value = 0.012 for weekday; chi-squared = 25.51, p-value = 0.0025 for weekend).

**Fig 4.** Duration of contacts by age category of participants during the weekday (Fig 4a) and weekend (Fig 4b) for all surveys conducted (2021-2023) in Fukuoka and Osaka prefectures indicated by each color. The mean and 95% confidence intervals are obtained by bootstrapping.

**Fig 4a.**
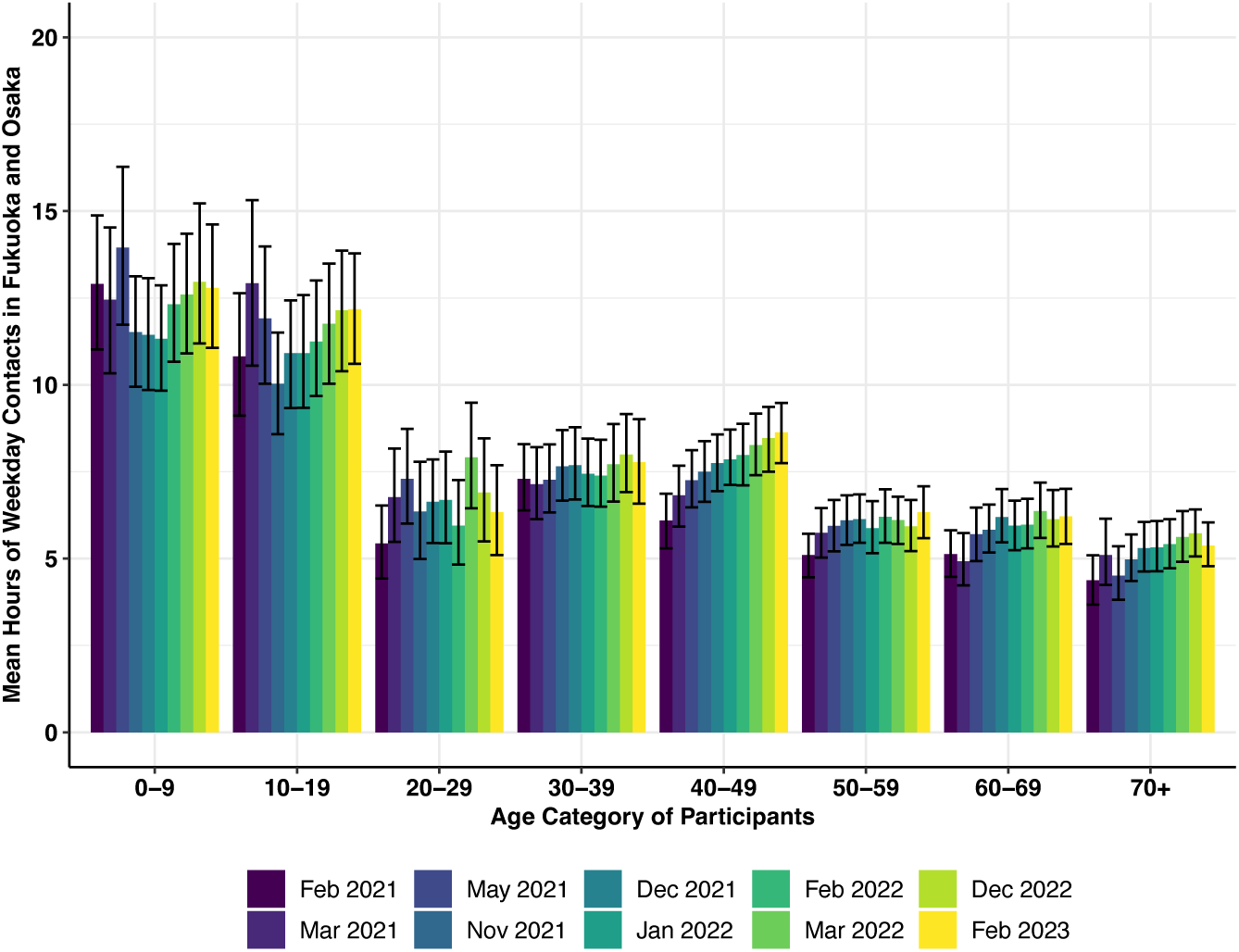
Weekday Contacts 2021-2023

**Fig 4b.**
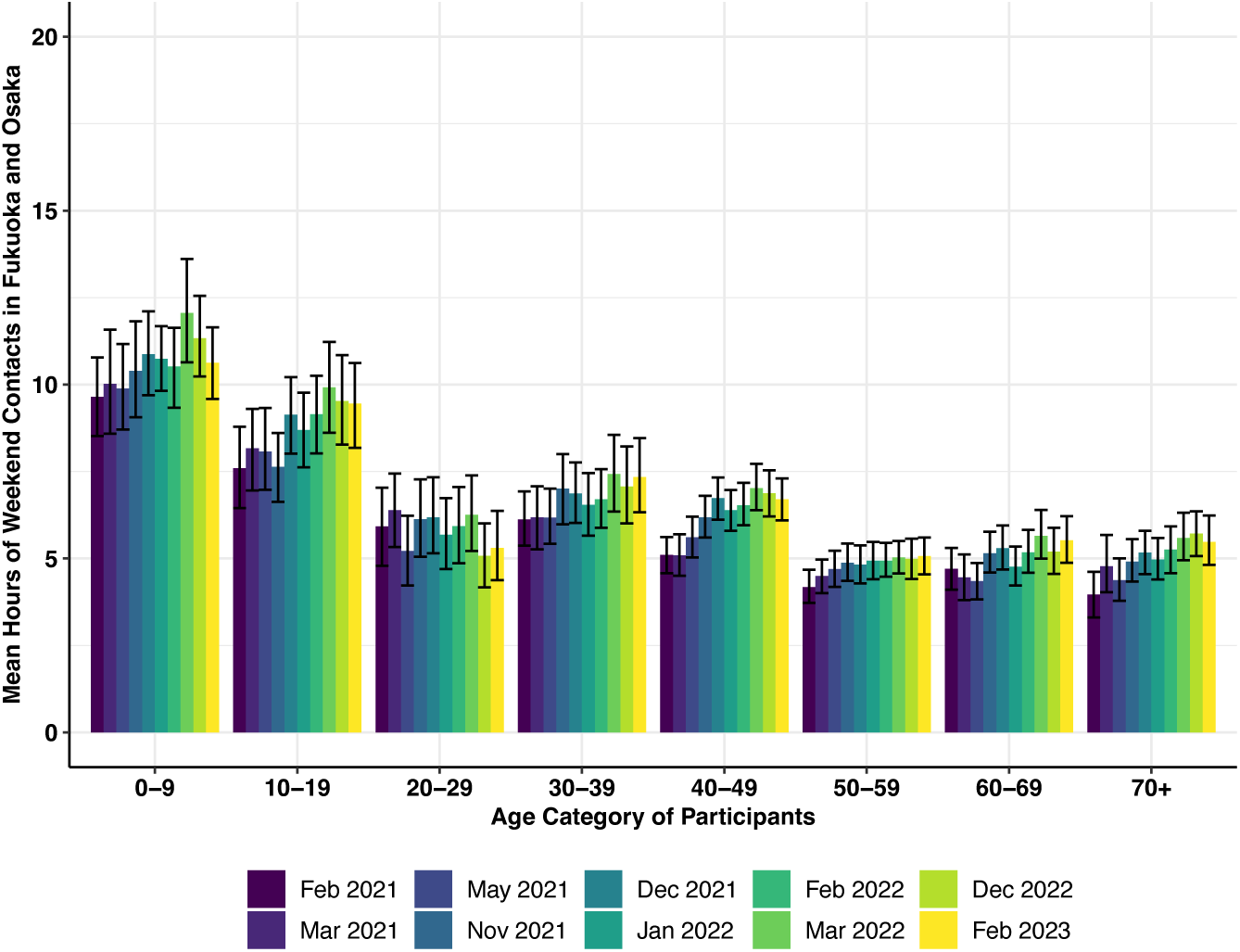
Weekend Contacts 2021-2023

### Association of Emergency Declarations (ED) and Semi-ED

We explored whether EDs (observed during surveys 1 and 3) were associated with reported contacts and their duration, and whether the strength of ED provided further granularity. After adjusting for age and sex in a generalized linear model, the issuance of an ED was negatively associated with the mean number of contacts during weekends (Adjusted CRR: 0.84 (95% CI: 0.79-0.88) compared with periods without any ED, yet there was no association during weekdays (Adjusted CRR: 0.96 (95% CI: 0.91-1.01). On the other hand, there was a slightly positive association between the issuance of a semi-ED (surveys 7 and 8) with contacts during weekends (Adjusted CRR: 1.06 (95% CI: 1.01-1.12)) and weekdays (Adjusted CRR: 1.05 (95% CI: 1.00-1.11)) compared with periods without any ED.

For example, a woman in her 40’s would contact an average of 3.32 individuals (95% CI: 3.09-3.56) during weekends when an ED was issued, but she would contact an average of 3.97 individuals (95% CI: 3.74-4.20) during periods when any level of ED was absent (Table 2). Since surveys 9 and 10 were conducted two years after the initial survey and could have significantly different contact patterns due to “pandemic fatigue,” the same analysis was conducted by excluding these two surveys, but the negative association between ED and contacts remained the same.

**Table 2:** Predicted frequency and duration of contacts for a woman in her 40’s during the weekday or weekend. Each row is based on a multivariable regression model that included age, sex, and the level of Emergency Declaration (ED) that was either absent, semi-ED or full ED. Each model includes all 10 survey time points from February 2021 to February 2023 (N=16,178 individuals).

|  | Scenario: Emergency Declaration (ED) absent<br>(Surveys 2,4,5,6,9, and 10) | Scenario: Period with Semi-ED<br>(Surveys 7 and 8) | Scenario: Period with ED<br>(Surveys 1 and 3) |
| --- | --- | --- | --- |
| **Outcome 1: Predicted Frequency of Contacts per individual (95% CI)** |  |  |  |
| a) Weekday | 5.79 (5.46-6.12) | 6.08 (5.68-6.52) | 5.53 (5.17-5.92) |
| b) Weekend | 3.97 (3.74-4.20) | 4.22 (3.93-4.52) | 3.32 (3.09-3.56) |
| **Outcome 2: Predicted Duration of Contacts (hours) per individual (95% CI)** |  |  |  |
| a) Weekday | 6.91 (6.55-7.28) | 7.13 (6.69-7.60) | 6.24 (5.86-6.65) |
| b) Weekend | 5.86 (5.55-6.18) | 6.20 (5.80-6.62) | 5.09 (4.76-5.43) |
| **Outcome 3: Predicted Contacts at Restaurants and Bars per individual (95% CI)** |  |  |  |
| a) Weekday | 0.022 (0.021-0.024) | 0.020 (0.019-0.022) | 0.017 (0.016-0.019) |
| b) Weekend | 0.035 (0.032-0.038) | 0.034 (0.031-0.037) | 0.024 (0.022-0.026) |

When surveys 1 and 3 (ED issued) were compared with survey 2 (ED absent) after adjusting for age and sex, there was no evidence of an association between the frequency of contacts and the issuance of an ED during the weekday (Adjusted CRR: 1.03 (95% CI: 0.95-1.12) or the weekend (Adjusted CRR: 0.95 (95% CI: 0.88-1.04).

The issuance of an ED was negatively associated with the duration of contacts during both weekdays (Adjusted CRR: 0.90 (95% CI: 0.86-0.95)) and weekends (Adjusted CRR: 0.87 (95% CI: 0.82-0.91)) after adjusting for age and sex. Following the same example given previously, a woman in her 40’s would have an average contact duration of 5.09 hours (95% CI: 4.76-5.43) with other individuals during weekends when an ED was issued and 5.86 hours (95%CI: 5.55-6.18 during periods without any ED.

Because one of the key restrictions during ED was either complete closure or shortening of restaurant/bar hours with restricted hours of serving alcohol (Supp. Table 2), we investigated the number of individuals who reported contacts at restaurants/bars. For this analysis, the contact location of restaurants and bars were combined which included izakaya—informal Japanese bars where alcohol and food are served—karaoke, and movie theatres. After adjusting for age and sex, these contacts were negatively associated with the issuance of ED during weekdays (Adjusted CRR: 0.77 (95% CI: 0.72-0.83)) and weekends (Adjusted CRR: 0.68 (95% CI: 0.63-0.74)) compared to periods when ED was absent.

### Factors associated with contact patterns

Based on our multivariable regression model, an individual with the reference characteristics had an expected 2.35 contacts (95% CI: 1.70-3.23) during the weekday. Each variable was compared to this reference, and we reported on characteristics that were statistically different to this value (Table 3). To check the model fit, the predicted values of weekday contacts were plotted against the observed values reported from the February 2023 contact survey (Figure 2). Residual plots were also evaluated (Supp. Fig 5). Heteroscedasticity was evident (Supp. Fig 5 a-b) which was expected due to the sample distribution showing overdispersion. Model fit was good as the error terms of each predictor in the model showed approximate linearity (Supp. Fig 5 c-f).

**Table 3:** Characteristics of participants and their reported number of weekday contacts from the February 2023 contact survey conducted in Fukuoka and Osaka prefectures compared to the reference category. The relative mean number of contacts per day is indicated as the contact rate ratio (CRR). The crude CRR is from a univariate regression model and the adjusted CRR is from a multivariable regression model. Rows highlighted in yellow are the variables that were associated (p<0.05) with the mean number of contacts in the multivariable regression model.

| Category | Number of Participants | % of Participants | Mean Number of Contacts | Median Number of Contacts | Lower IQR (Contacts) | Upper IQR (Contacts) | Crude Contact Rate Ratio | Lower 95% CI (Crude) | Upper 95% CI (Crude) | Adjusted Contact Rate Ratio | Lower 95% CI (Adjusted) | Upper 95% CI (Adjusted) |
| --- | --- | --- | --- | --- | --- | --- | --- | --- | --- | --- | --- | --- |
| **Participant Age (years)** |  |  |  |  |  |  |  |  |  |  |  |  |
| 0-5 | 45 | 2.87 | 13.27 | 5.00 | 2.00 | 14.00 | 1.56 | 1.01 | 2.40 | 1.46 | 0.37 | 5.78 |
| 6-17 | 136 | 8.67 | 17.01 | 9.00 | 4.00 | 24.00 | 2.17 | 1.64 | 2.89 | 1.25 | 0.49 | 3.16 |
| 18-29 | 120 | 7.65 | 10.53 | 4.00 | 2.00 | 8.00 | 1.20 | 0.90 | 1.62 | 1.00 | 0.74 | 1.36 |
| 30-39 | 162 | 10.33 | 7.19 | 3.00 | 2.00 | 6.00 | 0.82 | 0.63 | 1.07 | 0.81 | 0.65 | 1.02 |
| 40-49 (reference) | 275 | 17.53 | 8.46 | 4.00 | 2.00 | 7.00 | NA | NA | NA | NA | NA | NA |
| 50-59 | 331 | 21.10 | 6.03 | 3.00 | 1.00 | 6.00 | 0.70 | 0.56 | 0.87 | 0.88 | 0.74 | 1.06 |
| 60+ | 500 | 31.87 | 6.05 | 3.00 | 1.00 | 5.00 | 0.70 | 0.57 | 0.86 | 1.07 | 0.88 | 1.29 |
| **Number in Household** |  |  |  |  |  |  |  |  |  |  |  |  |
| 1 person | 154 | 9.82 | 3.45 | 2.00 | 0.00 | 3.00 | 0.51 | 0.40 | 0.66 | 1.36 | 1.03 | 1.82 |
| 2 people | 378 | 24.09 | 5.88 | 2.00 | 1.00 | 5.00 | 0.90 | 0.74 | 1.09 | 1.09 | 0.92 | 1.28 |
| 3 people (reference) | 408 | 26.00 | 6.00 | 3.00 | 2.00 | 6.00 | NA | NA | NA | NA | NA | NA |
| 4 people | 368 | 23.45 | 10.88 | 5.00 | 3.00 | 8.00 | 1.77 | 1.46 | 2.14 | 1.46 | 1.24 | 1.73 |
| 5+ people | 261 | 16.63 | 13.34 | 5.00 | 3.00 | 10.00 | 2.03 | 1.64 | 2.51 | 1.68 | 1.40 | 2.02 |
| **Occupation** |  |  |  |  |  |  |  |  |  |  |  |  |
| Company Director/Executive Manager | 17 | 1.08 | 5.00 | 3.00 | 1.00 | 6.00 | 0.68 | 0.36 | 1.29 | 0.95 | 0.54 | 1.65 |
| Company employee (reference) | 445 | 28.36 | 7.77 | 4.00 | 2.00 | 8.00 | NA | NA | NA | NA | NA | NA |
| Temporary contractor | 81 | 5.16 | 9.46 | 3.00 | 1.00 | 6.00 | 1.05 | 0.77 | 1.43 | 0.79 | 0.60 | 1.03 |
| Government employee | 45 | 2.87 | 17.02 | 5.00 | 3.00 | 15.00 | 2.17 | 1.45 | 3.26 | 1.50 | 1.05 | 2.13 |
| Commerce industry/independent business | 59 | 3.76 | 4.36 | 3.00 | 2.00 | 4.00 | 0.60 | 0.42 | 0.87 | 0.72 | 0.53 | 0.98 |
| Non-healthcare professional (e.g. lawyer, accountant) | 12 | 0.76 | 11.00 | 4.00 | 2.00 | 10.00 | 1.45 | 0.68 | 3.08 | 1.16 | 0.61 | 2.22 |
| Healthcare professional | 19 | 1.21 | 16.21 | 8.00 | 6.00 | 16.00 | 2.22 | 1.21 | 4.08 | 2.27 | 1.35 | 3.80 |
| Social work service (e.g. long-term care facility) | 3 | 0.19 | 15.33 | 14.00 | 13.00 | 17.00 | 2.42 | 0.54 | 10.84 | 2.45 | 0.70 | 8.59 |
| Part-time job (non-homemaker) | 106 | 6.76 | 8.92 | 4.00 | 1.00 | 8.00 | 1.13 | 0.85 | 1.50 | 1.15 | 0.90 | 1.47 |
| Homemaker (with part-time job) | 57 | 3.63 | 6.18 | 4.00 | 3.00 | 6.00 | 0.87 | 0.61 | 1.26 | 0.80 | 0.58 | 1.10 |
| Homemaker (without part-time job) | 200 | 12.75 | 4.40 | 3.00 | 1.00 | 4.00 | 0.59 | 0.47 | 0.73 | 0.80 | 0.55 | 1.15 |
| Freelancer | 36 | 2.29 | 8.08 | 3.00 | 1.00 | 4.00 | 0.91 | 0.58 | 1.43 | 1.49 | 1.00 | 2.23 |
| Infant (pre-school) | 24 | 1.53 | 6.21 | 4.00 | 2.00 | 6.00 | 0.85 | 0.50 | 1.47 | 1.05 | 0.27 | 4.13 |
| Kindergartner | 26 | 1.66 | 21.58 | 16.00 | 4.00 | 30.00 | 2.93 | 1.74 | 4.95 | 1.31 | 0.33 | 5.15 |
| Primary school student | 60 | 3.82 | 18.02 | 10.00 | 4.00 | 33.00 | 2.58 | 1.80 | 3.68 | 0.83 | 0.32 | 2.17 |
| Middle school student | 45 | 2.87 | 17.51 | 8.00 | 4.00 | 15.00 | 2.30 | 1.53 | 3.45 | 0.70 | 0.27 | 1.84 |
| High school student | 30 | 1.91 | 12.40 | 5.50 | 2.00 | 18.00 | 1.68 | 1.03 | 2.74 | 0.60 | 0.25 | 1.41 |
| Post high school prep student | 1 | 0.06 | 4.00 | 4.00 | 4.00 | 4.00 | 0.64 | 0.05 | 8.50 | 0.65 | 0.07 | 5.80 |
| College student | 46 | 2.93 | 11.11 | 3.50 | 2.00 | 8.00 | 1.35 | 0.91 | 2.02 | 0.84 | 0.50 | 1.42 |
| Unemployed/helps at the house | 72 | 4.59 | 3.01 | 2.00 | 1.00 | 3.00 | 0.40 | 0.28 | 0.55 | 0.55 | 0.36 | 0.85 |
| Pension retiree | 179 | 11.41 | 3.78 | 2.00 | 1.00 | 4.00 | 0.51 | 0.41 | 0.64 | 0.56 | 0.38 | 0.83 |
| Other | 6 | 0.38 | 6.67 | 3.50 | 0.00 | 5.00 | 0.83 | 0.29 | 2.40 | 0.74 | 0.29 | 1.88 |
| **Moving Prefectures for Work/School during the Month** |  |  |  |  |  |  |  |  |  |  |  |  |
| Less than 6 times (reference) | 1465 | 93.37 | 7.54 | 3.00 | 2.00 | 7.00 | NA | NA | NA | NA | NA | NA |
| More than 6 times | 104 | 6.63 | 15.81 | 5.00 | 3.00 | 10.00 | 1.99 | 1.51 | 2.63 | 1.44 | 1.14 | 1.82 |
| **Location of Work** |  |  |  |  |  |  |  |  |  |  |  |  |
| Telework at home | 320 | 20.40 | 3.56 | 2.00 | 1.00 | 4.00 | 0.37 | 0.31 | 0.44 | 0.57 | 0.46 | 0.71 |
| Workplace (reference) | 610 | 38.88 | 10.38 | 5.00 | 3.00 | 9.00 | NA | NA | NA | NA | NA | NA |
| School | 188 | 11.98 | 17.70 | 8.00 | 3.00 | 24.00 | 1.79 | 1.45 | 2.21 | 0.81 | 0.52 | 1.26 |
| Not Employed | 451 | 28.74 | 4.20 | 2.00 | 1.00 | 4.00 | 0.42 | 0.36 | 0.49 | 0.70 | 0.49 | 0.99 |
| **Frequency of Teleworking** |  |  |  |  |  |  |  |  |  |  |  |  |
| Never (reference) | 818 | 52.14 | 11.28 | 5.00 | 2.00 | 9.00 | NA | NA | NA | NA | NA | NA |
| Few times per month | 85 | 5.42 | 6.96 | 5.00 | 3.00 | 8.00 | 0.69 | 0.51 | 0.93 | 0.76 | 0.59 | 0.98 |
| 2-3 times per week | 96 | 6.12 | 6.96 | 3.00 | 2.00 | 6.00 | 0.59 | 0.45 | 0.78 | 0.81 | 0.62 | 1.06 |
| 4-5 times per week | 119 | 7.58 | 2.58 | 2.00 | 1.00 | 3.00 | 0.26 | 0.20 | 0.33 | 0.66 | 0.50 | 0.87 |
| **Reported Location of Contact** |  |  |  |  |  |  |  |  |  |  |  |  |
| No Contact at Home (reference) | 279 | 17.78 | 3.99 | 1.00 | 0.00 | 3.00 | NA | NA | NA | NA | NA | NA |
| Home | 1290 | 82.22 | 8.97 | 4.00 | 2.00 | 8.00 | 2.86 | 2.38 | 3.44 | 2.65 | 2.16 | 3.26 |
| No Contact at Others' Home (reference) | 1517 | 96.69 | 7.96 | 3.00 | 2.00 | 7.00 | NA | NA | NA | NA | NA | NA |
| Others' Home | 52 | 3.31 | 11.83 | 6.00 | 3.00 | 8.00 | 1.57 | 1.06 | 2.33 | 1.30 | 0.95 | 1.79 |
| No Contact at School (reference) | 1400 | 89.23 | 6.42 | 3.00 | 2.00 | 6.00 | NA | NA | NA | NA | NA | NA |
| School | 169 | 10.77 | 21.92 | 10.00 | 6.00 | 30.00 | 3.70 | 2.99 | 4.59 | 3.51 | 2.50 | 4.93 |
| No Contact at Restaurant (reference) | 1479 | 94.26 | 7.81 | 3.00 | 2.00 | 7.00 | NA | NA | NA | NA | NA | NA |
| Restaurant | 90 | 5.74 | 12.57 | 6.00 | 4.00 | 10.00 | 1.72 | 1.28 | 2.33 | 1.34 | 1.03 | 1.75 |
| No Contact at Bar (reference) | 1536 | 97.90 | 7.89 | 3.00 | 2.00 | 7.00 | NA | NA | NA | NA | NA | NA |
| Bar | 33 | 2.10 | 17.00 | 9.00 | 4.00 | 22.00 | 2.38 | 1.46 | 3.88 | 2.18 | 1.42 | 3.36 |
| **Frequency of Mask Wearing during the Day** |  |  |  |  |  |  |  |  |  |  |  |  |
| <3 hrs (reference) | 798 | 50.86 | 4.40 | 3.00 | 1.00 | 5.00 | NA | NA | NA | NA | NA | NA |
| 3-6 hrs | 273 | 17.40 | 7.48 | 3.00 | 2.00 | 7.00 | 1.61 | 1.35 | 1.92 | 1.18 | 1.00 | 1.39 |
| 6-9 hrs | 258 | 16.44 | 8.77 | 5.00 | 2.00 | 9.00 | 2.01 | 1.68 | 2.40 | 1.27 | 1.07 | 1.51 |
| 9-12 hrs | 149 | 9.50 | 18.64 | 7.00 | 4.00 | 20.00 | 4.06 | 3.25 | 5.08 | 2.14 | 1.71 | 2.68 |
| 12+ hrs | 91 | 5.80 | 22.97 | 6.00 | 3.00 | 22.00 | 4.69 | 3.55 | 6.19 | 2.53 | 1.92 | 3.34 |
| **Frequency of Handwashing during the Past 3 hrs** |  |  |  |  |  |  |  |  |  |  |  |  |
| None | 258 | 16.44 | 6.24 | 3.00 | 1.00 | 7.00 | 0.72 | 0.58 | 0.89 | 0.74 | 0.62 | 0.88 |
| 1 time (reference) | 532 | 33.91 | 8.70 | 4.00 | 2.00 | 8.00 | NA | NA | NA | NA | NA | NA |
| 2 times | 311 | 19.82 | 9.14 | 3.00 | 2.00 | 7.00 | 1.03 | 0.84 | 1.26 | 0.98 | 0.83 | 1.15 |
| 3 times | 213 | 13.58 | 7.20 | 3.00 | 2.00 | 6.00 | 0.85 | 0.68 | 1.06 | 0.74 | 0.62 | 0.89 |
| 4 times | 76 | 4.84 | 8.53 | 4.00 | 2.00 | 8.00 | 1.00 | 0.71 | 1.40 | 0.77 | 0.59 | 1.02 |
| 5 times | 92 | 5.86 | 9.04 | 3.00 | 1.00 | 8.00 | 1.00 | 0.73 | 1.37 | 1.04 | 0.81 | 1.34 |
| 6+ times | 87 | 5.54 | 6.85 | 3.00 | 1.00 | 6.00 | 0.80 | 0.58 | 1.10 | 0.67 | 0.52 | 0.87 |
| **Number of Times Vaccinated against COVID-19** |  |  |  |  |  |  |  |  |  |  |  |  |
| Never Vaccinated | 264 | 16.83 | 11.69 | 3.00 | 2.00 | 9.00 | 1.54 | 1.27 | 1.86 | 1.06 | 0.88 | 1.28 |
| 1 dose | 11 | 0.70 | 4.00 | 4.00 | 2.00 | 6.00 | 0.63 | 0.27 | 1.45 | 0.59 | 0.31 | 1.16 |
| 2 doses | 143 | 9.11 | 7.05 | 4.00 | 2.00 | 7.00 | 1.01 | 0.79 | 1.29 | 0.88 | 0.71 | 1.08 |
| 3+ doses (reference) | 1136 | 72.40 | 7.42 | 3.00 | 2.00 | 7.00 | NA | NA | NA | NA | NA | NA |
| **Perspective on COVID-19** |  |  |  |  |  |  |  |  |  |  |  |  |
| 1 - Most concerned (reference) | 344 | 21.92 | 8.06 | 3.00 | 2.00 | 6.00 | NA | NA | NA | NA | NA | NA |
| 2 - Concerned | 562 | 35.82 | 6.87 | 3.00 | 2.00 | 6.00 | 0.90 | 0.74 | 1.08 | 1.05 | 0.90 | 1.23 |
| 3 - Neutral | 186 | 11.85 | 7.17 | 3.00 | 1.00 | 7.00 | 0.94 | 0.73 | 1.20 | 0.99 | 0.80 | 1.22 |
| 4 - Not concerned | 183 | 11.66 | 6.67 | 3.00 | 2.00 | 6.00 | 0.93 | 0.72 | 1.19 | 1.08 | 0.88 | 1.34 |
| 5 - Least concerned | 71 | 4.53 | 4.89 | 2.00 | 1.00 | 4.00 | 0.65 | 0.45 | 0.93 | 0.73 | 0.54 | 1.00 |
| 6 - Do not know | 223 | 14.21 | 14.14 | 6.00 | 3.00 | 18.00 | 1.97 | 1.55 | 2.49 | 0.60 | 0.39 | 0.92 |

Contact patterns varied by location (Table 3). Participants who reported contacts at home, school, restaurants, and bar settings during the weekday had higher contacts than those who did not report any contacts at these locations. Those who lived in a household of four or five people had higher contacts than those who lived in a household of three people. The association between contacts and occupation differed depending on the occupation type where healthcare professionals and government employees, including public school teachers, had significantly higher contacts. Different work conditions were associated with contact patterns. While those who teleworked had lower contacts, those who moved to a different prefecture at least six times in the past month for work/school purposes had higher contacts. As of February 2023, among those who reported as employed or attending school, 73.2% (818/1118) reported they have never teleworked from home. Disease mitigation measures, such as mask wearing frequency increased significantly with higher number of contacts, but there was no increasing trend of handwashing frequency with more contacts. There was no evidence of association between contacts and vaccination status and their level of concern in getting infected with COVID-19. Note that among those who reported their frequency of teleworking, there were 451 people (28.71%) who reported not employed and there were 15 people (0.96%) who reported NA on their vaccination status (not shown on Table 3).

## Discussion

Our study is one of the few that have characterized social contact patterns in Japan during the COVID-19 pandemic through repeated cross-sectional surveys. Contact surveys were conducted in Japan during and after the Tokyo Olympics in August 2021 that resulted in having a median of three contacts per day (mean of 8.92) (23), similar to our results from the March 2022 survey.

Although our surveys did not capture contact patterns in 2020, a study using mobility data has shown that there was a 70% reduction in daily total contacts in April 2020 after there were government recommendations to telework and close the public schools (24). This is comparable to countries such as Norway (67-73% decrease) (25), France (70% decrease) (26), Germany (73% decrease) (27), United Kingdom (74% decrease) (17), Belgium (80% decrease) (28)and United States (82% decrease) (29) where strict lockdowns were implemented. Many contact patterns remained subdued even after post-lockdowns (e.g. China, 21 European countries) (30,31).

Although Japan never implemented lockdowns, our results suggest that the objective of ED issuance was met as it was associated with changes in frequency and duration of contacts. Particularly during the weekend, there were fewer contacts and shorter duration of contacts. This suggests that individuals may have changed their behavior during the days when they have more control over compared to the weekdays when they either commute to school or work. Even as of February 2023, two years after the second ED, the majority (73.2%) of those who were employed or attending school had never worked from home. This corresponds to a survey that showed among approximately 20,000 employed individuals, 70.3% were not teleworking during the first ED in April 2020 when stay-at-home and teleworking recommendations were issued for the first time during the pandemic (32). Prior to the pandemic, teleworking was a practice rarely implemented in Japanese society; a national survey in September 2019 showed 20.2% of 2,118 companies had implemented teleworking strategies for their employees (33).

The duration of contacts increased with time particularly among children, teenagers, and adults in their 40s. This is correlated with governmental restrictions that were gradually lifted, such as having less stringent measures during semi-EDs compared to EDs. However, the mean number of contacts remained subdued across the first three surveys in 2021, including March when ED was absent, showing that the lifting of ED was not what may have influenced the increase in contact patterns per se. These can be signs of pandemic fatigue where there may be a gradual shift towards behavior more like pre-pandemic times. It is also worth noting that there were other factors that also increased with time, such as vaccination and circulation of omicron, a less virulent strain of SARS-CoV-2. It is, however, important to note that even after ED was lifted in March 2021 before mass vaccination started, the frequency and duration of contacts did not change. This contrasts with periods when the first and second lockdowns were lifted in the UK, also prior to vaccine rollout to the entire public, adults (18 years and above) increased their contacts from 29 to 59% compared to during the first lockdown (19). These empirical data illustrate how contact patterns can vary from country to country during a pandemic.

Understanding how individuals behave, especially during periods prior to vaccine rollout, may provide hints on the level of necessity, timing, and severity of governmental recommendations in future epidemics.

The Japanese population that had consistently low number of contacts was those over 70, which consist of 22.6% of the national population (15). They not only met with fewer than five contacts during the week, but each contact remained short (average of one hour of contact per day) with very limited physical contact throughout the pandemic (Supp. Fig 3a). Their contact patterns also did not change after mass vaccination started from May 2021 for 65 years and above. With the combination of shielding the older populations in long-term care facilities (34), this may have helped in preventing a surge of COVID-19 outbreaks and deaths amongst this population during the beginning of the pandemic—an issue that was apparent in the US (35) and the UK (35,36).

“Prosocial behavior,” such as physical distancing, mask wearing, and getting tested as described by Sachs JD et al., (3), is critical in pandemic control, and countries in WHO’s Western Pacific region were quick in encouraging it as part of their “suppression strategy” (3). Our results in Japan reflect this. In addition to EDs, the 3C policy was implemented from early 2020 which was especially relevant in the Japanese context as our results showed that indoor contacts were more frequent than outdoor contacts (Supp. Fig 4a and 4b). Individuals who went to restaurants and bars had higher contacts compared to those who did not. The close contact that is likely in these settings highlight the importance of mitigation approaches such as improved ventilation for disease control (37).

In our surveys there were increased hours of mask wearing with higher number of contacts, demonstrating prosocial behavior. Frequency of mask wearing continued to be stable throughout 2021 to 2023 which contrasts with the UK where mask wearing was strongly associated with changes in government policy (19). On the other hand, handwashing was not associated with the frequency of contacts. This may be due to the public message of 3C where avoiding crowded conditions and close-contact settings were highlighted in Japan more than handwashing and disinfection—another key difference compared to other countries (5). Contacts were not associated with vaccination status, opposite from the European countries where vaccinated individuals reported higher contacts (38) This could be due to Japan not implementing any restrictions in accessing public or indoor areas due to vaccination status, unlike in Europe where vaccine certificates were issued. Such differences suggest behavioral patterns that are triggered by risk perception vs. governmental intervention. The number of deaths increased after the introduction of the omicron variant, but the suppression strategy in Japan from 2020-2021 – a combination of reduced contacts, prosocial behavior, and high vaccination coverage – may have helped in keeping a low cumulative mortality like other Western Pacific countries where the suppression strategy was implemented early (3).

There are limitations of our study. Our surveys were limited to Osaka and Fukuoka prefectures, where incidence has been typically higher than the remaining 45 prefectures in Japan. Extrapolating our findings to other prefectures in Japan should be done with caution, as contact rates may be lower. However, Japanese contact survey studies prior to the COVID-19 pandemic by Ibuka et al. (12) and Munasinghe et al. (13) did not report any differences in contact rates across prefectures, while Tsuzuki et al. (23) examined contact rates between rural and urban areas and did not find any meaningful differences. We can therefore have moderate confidence in the relevance of our findings for the rest of the country. Selection bias could have been introduced as the surveys captured a sample population that had a slightly higher vaccination coverage than the national average. Recording contacts retrospectively may result in recall bias, leading to an underestimation of contacts. Lastly, due to the limited number of questions that could be asked in each survey, our multivariable regression model could have included covariates that were not included in the model, leading to residual bias. Some of these variables could have impacted contact patterns with time. Thus, future work is planned to investigate factors that could have influenced the change in contacts during the pandemic by analyzing the same individuals across multiple time points.

Compared to other countries, Japan has been unique in tackling the COVID-19 pandemic without implementing lockdowns and long school closures; it mostly relied on recommendations that were not legal enforcements. Prosocial behavior, such as limiting contacts especially during the weekend in 2021 and continued mask wearing until 2023 were evident. Although issuance of EDs occurred as the pandemic progressed in waves, contacts remained subdued compared to pre-pandemic times. By recognizing some of the key factors that influence contact patterns in Japan, it can contextualize mathematical models of SARS-CoV-2 that are developed to understand the transmission dynamics and disease. Behavior can be difficult to predict and contact patterns may differ in a future pandemic. The age-specific contacts from our study can be utilized in mathematical modelling, for example to characterize infectious disease spread and support pandemic planning. Most importantly, identifying these country-specific factors that influence human behavior provides further support for policies in controlling disease transmission that are context specific, not only for COVID-19 but also for other infectious diseases that may emerge in the future.

## Conclusions

We conducted repeated surveys in Japan to understand how the population modified its behavior and contact patterns during the pandemic. Our results showed that daily contacts dropped by approximately 50% during the pandemic, rebounding only slowly after the government recommendations were relaxed. People proceeded with caution as they wore masks longer if they had more contacts and consistently wore masks until 2023, even after a full year since the last governmental recommendation was lifted. The frequency of contacts did not increase after individuals received the COVID-19 vaccine which is in contrast with European countries where the opposite trend was reported. Our findings provide evidence on the importance of reduced contacts, careful behavior, and high vaccination coverage that potentially limited disease transmission and mortality in Japan.

## Data Availability

All accompanying code and data to generate all the figures in the main manuscript is available on GitHub: https://github.com/tomokanakamura/jp_contactsurvey.git

## List of abbreviations

AIC: Akaike Information Criterion
CI: confidence interval
COVID-19: coronavirus disease 2019
CRR: Contact Rate Ratio
ED: Emergency Declaration

## Ethics

Participation in the survey was voluntary and all analyses were performed on anonymized data. The study design including the survey questions and informed consent were approved by the ethics committee of Nagasaki University School of Tropical Medicine and Global Health (approval number: NU_TMGH_2022_162_4). Informed consent was obtained from all participants who provided data from the surveys.

## Data availability

Aggregated datasets are available to reproduce all the figures in the main manuscript. A processed dataset with the selected covariates in the multivariable generalized linear model is available to reproduce Table 3. Due to potentially sensitive and identifiable information included, the original dataset of the contact surveys is not made public and is available from the corresponding author upon reasonable request.

## Code availability

All accompanying code to generate all the figures in the main manuscript is available on GitHub: https://github.com/tomokanakamura/jp_contactsurvey.git

## Acknowledgements

We would like to acknowledge F-press that implemented the social contact surveys online and the study population in Osaka and Fukuoka prefectures who kindly participated in our surveys. We thank John Edmunds and our colleagues within the Centre for Mathematical Modelling of Infectious Diseases at LSHTM, the National Institute for Infectious Diseases in Japan, and Nagasaki University’s School of Tropical Medicine and Global Health and the Department of Clinical Medicine for input during the development of this study.

## Author Contributions

TN, KA, KMO, RK, and AE contributed to designing the contact surveys. TN and RK collected the contact survey data. TN processed and analyzed the contact survey data. TN and KMO designed the statistical analysis. KA, KMO, KEA, MS, AE, YI, and HO provided feedback on the study design and analysis plan. All authors contributed to the interpretation of the study results, writing and critical revision of the manuscript.

